# Metabolomic signatures of hypocaloric dietary interventions associate with breast cancer risk in the Nurses’ Health Study II

**DOI:** 10.1101/2025.08.18.25333805

**Authors:** Annalise Schweickart, Cheng Peng, Oana Zeleznik, William Dartora, Maurice A. Hurd, Alex Buga, Madison L. Kackley, Clary Clish, Laura Beth McIntire, Jeff S. Volek, Vicky Makker, A. Heather Eliassen, Marcus D. Goncalves, Rulla M. Tamimi, Jan Krumsiek

## Abstract

**Background:** An individual’s metabolic state plays a critical role in breast cancer (BC) risk, influenced by factors such as obesity and insulin signaling. Hypocaloric diets induce metabolic changes that influence these metabolic factors, thereby potentially influencing BC risk. However, it remains unclear whether metabolic profiles like those induced by such beneficial diets are associated with BC risk.

**Methods:** We compared the impact of a hypocaloric low-carbohydrate ketogenic diet (KD) and a low-fat diet (LFD) on BC risk in two stages. First, we developed metabolomics-based scores representing the metabolic states resulting from these two hypocaloric diets. Plasma metabolomics data of 43 individuals from two controlled dietary interventions were analyzed (N = 31 KD, N = 12 LFD) and a metabolite-based score was generated for both KD and LFD using diet-induced fold-changes. Second, these scores were applied to metabolomics data from a nested case-control study of participants from the Nurses’ Health Study II (NHSII, 1,058 BC cases, 1,054 controls, predominantly premenopausal women). Using multivariable-adjusted models, we assessed the association between the metabolomic scores and BC risk.

**Results:** KD and LFD had similar but distinct metabolic signatures. Both metabolomics scores were positively associated with breast cancer risk in NHSII. Women in the highest quartile of the KD metabolomic score had a 37% increased risk of BC compared to women in the lowest quartile (p=0.021). Similarly, women in the highest quartile of the LFD metabolomic score had a 32% increased BC risk compared to women in the lowest quartile (p=0.008). Similar increases in risk were seen when further adjusting for BMI at age 18 and weight change since age 18. Increased levels of cholesterol esters (CE), particularly CE 22:6, and long-chain polyunsaturated triglycerides were associated with higher risk in both diet scores, while increases in short-chain, more saturated triglycerides were associated with lower risk.

**Conclusion:** Metabolomic profiles resembling those induced by hypocaloric ketogenic and low-fat diets were unexpectedly associated with an increased risk of breast cancer in a predominantly premenopausal cohort. These associations were independent of BMI, highlighting the complex relationship between metabolic states and cancer risk, independent of actual dietary interventions.

## 1 Introduction

Breast cancer remains a major public health concern, with over 290,000 women diagnosed annually in the U.S. and nearly 44,000 deaths attributed to the disease^1^. Despite advancements in early detection, breast cancer incidence rates have been rising by 0.3% per year over the past two decades, highlighting a pressing need for more effective prevention strategies^2^.

One area of focus is the relationship between breast cancer risk and obesity, usually represented by the body mass index (BMI)^3–5^. This association varies depending on the stage of life: After menopause, high BMI is consistently associated with an increased risk of breast cancer. Tumorigenesis in this setting is thought to be driven by an expanded adipose tissue compartment, which drives hormonal changes and inflammation^6,7^. Interestingly, high BMI is correlated with a decreased risk of breast cancer in premenopausal women^7^. The mechanisms underlying this apparent protective effect remain largely unknown.

Beyond BMI, weight change can affect breast cancer risk; weight gain over short time periods has been linked to a higher risk of breast cancer in both premenopausal and postmenopausal women^8^. Encouragingly, evidence from the Nurses’ Health Study indicates that postmenopausal women who achieve sustained weight loss experience a reduced breast cancer risk^9^. The effects of weight loss on breast cancer risk in premenopausal women have not been conclusive^10^.

Related to BMI and weight change, insulin dynamics have also been implicated in breast cancer risk^11–13^. Multiple mouse models have connected insulin and hyperinsulinemia to the pathogenesis of breast cancer development through activation of the insulin receptor^14,15^. C-peptide, a marker of insulin secretion with a longer half-life than insulin itself, has been used in epidemiological studies to explore this relationship^16^. While findings have been inconsistent^17,18^, the largest study to date (N = 3,481) reported a positive association between C-peptide levels and invasive breast cancer risk in both pre- and post-menopausal cases^19^.

Given these links between BMI, weight gain, insulin secretion, and breast cancer, dietary interventions targeting these risk factors are of significant interest^11,20,21^. Dietary patterns contributing to hyperinsulinemia positively associate with breast cancer risk^21^, while high diet quality has been shown to correlate with a lower incident breast cancer risk^22^, indicating dietary intervention could reduce risk of breast cancer development. Caloric restriction through both hypocaloric ketogenic diets (KD) and low-fat diets (LFD) has proved effective as a weight loss strategy^23–25^, and in the case of KD, insulin-lowering as well^26–28^, which may hold promise for breast cancer risk reduction. The metabolic reprogramming of hypocaloric diets has been associated with increased insulin sensitivity, weight reduction, and improved lipid profiles, all of which could potentially modulate breast cancer risk^25^.

To improve our understanding of how hypocaloric diets with varying macronutrient compositions (high fat vs. low fat) influence breast cancer, we must first investigate the metabolic changes induced by caloric restriction and how those relate to breast cancer risk. Metabolomic profiling of blood allows us to evaluate the contribution of metabolites, circulating small molecules that reflect metabolic state, to various phenotypes, including dietary state^29–31,32^. While several studies have identified metabolites that are affected by KD and LFD dietary interventions^33–37^, none have evaluated these changes prospectively in human cohorts to ascertain breast cancer risk.

In this study, we developed metabolomic scores capturing the distinct metabolic states induced by hypocaloric ketogenic diet (KD) and low-fat diet (LFD) interventions in two controlled dietary trials (**Figure 1**). We then assessed whether these diet-induced metabolic profiles were associated with breast cancer risk in the predominantly premenopausal Nurses’ Health Study II.

**Figure 1.**
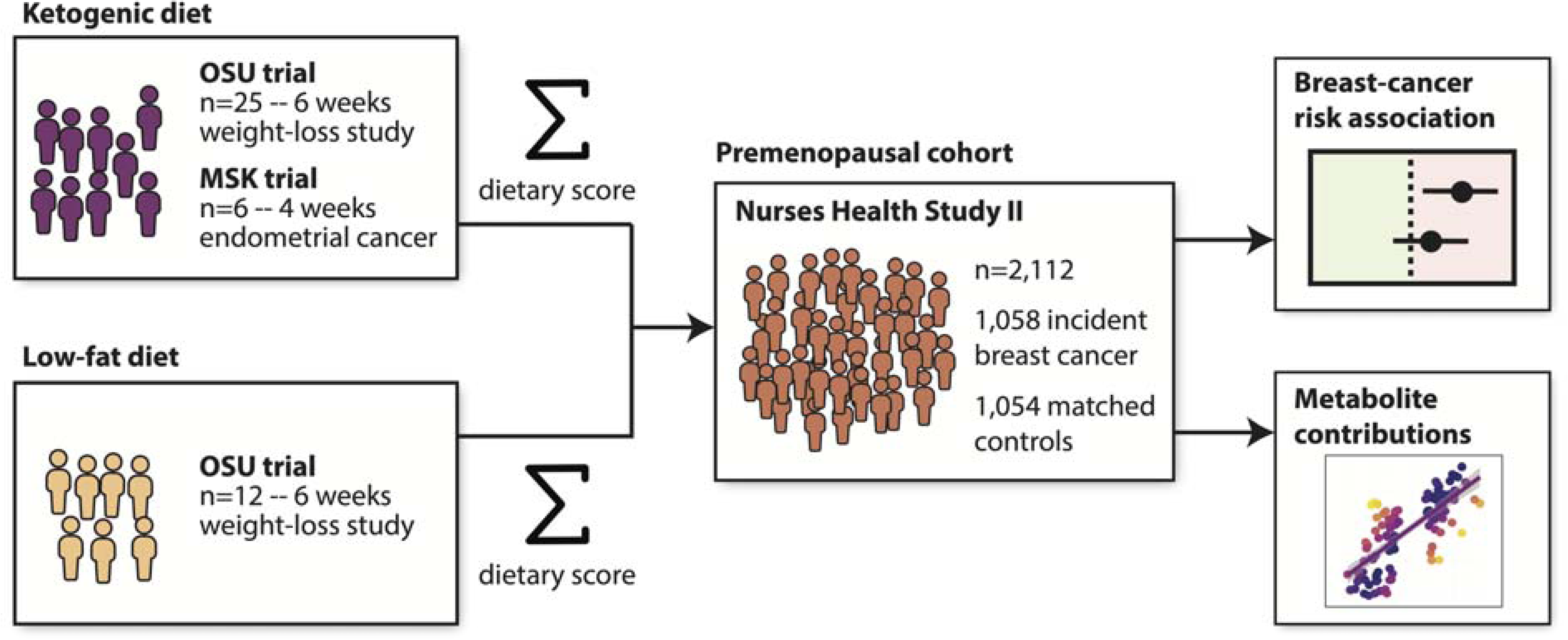
Study design. Metabolomic data from two controlled hypocaloric diet interventions (ketogenic and low-fat) were used to derive diet-specific scores, which were then applied to the NHSII nested case-control cohort to evaluate associations with breast cancer risk.

## 2 Methods

### 2.1 Study populations

#### OSU Cohort

This study was a prospective, placebo-controlled, double-blind trial evaluating a hypocaloric diet, where participants were selected based on eligibility criteria of BMI (27–35 kg/m²) and age (21–65 years)^38^. The study protocol was approved by The Ohio State University Institutional Review Board (IRB #: 2017H0395). All eligible participants signed an informed consent document approved by IRB before participating in the study. Stratified randomization was performed by dividing participants into groups based on age and BMI, categorized as either below or above the median of the inclusion range. After stratification, participants were assigned to one of two groups: a ketogenic diet (KD) plus ketone supplement group (N = 12, 6 men, 6 women) or a KD plus placebo group (N = 13, 6 men, 7 women). Additionally, a non-randomized group of participants matched by age and BMI received an isoenergetic and isonitrogenous low-fat diet (LFD) (N = 12, 6 men, 6 women) and underwent the same testing protocols as the KD groups. All meals for both the KD and LFD were prepared in a metabolic kitchen and provided 75% of estimated energy requirements for each participant. There were no significant differences in the baseline characteristics of those that completed the diets between the groups. All participants were Caucasian. Of the 25 individuals in the KD groups, the average weight loss was 8.3 ± 1.8%. In the LFD group, the average weight loss was 6.8 ± 3%. Information regarding exclusion criteria has been previously published^38^. Dietary composition can be found in **Supplementary Table 1**.

#### MSK Cohort

This was a multicenter, parallel, prospective, randomized, controlled feasibility study in treatment-naïve women with endometrial cancer during the presurgical window period between diagnosis and surgical staging. Participants were enrolled between February 2018 and August 2021 at seven Memorial Sloan Kettering (MSK) locations in New York and New Jersey (IRB #:17-396 A(14)). The 19 participants who fulfilled the enrollment criteria were randomized in a 2:1 fashion to receive a KD or to continue with their usual, standard diet (SD) until the day of surgery, 3-4 weeks later. Participants were predominantly white (60%), non-Hispanic (66%), middle-aged (median age 57, range 37-74), and obese (median BMI 37 with range 26-60 kg/m^2^). Of the 19 participants, 15 completed the study. The average weight loss was 5.5 ± 0.8%. Out of those 15 participants, 11 underwent metabolomic profiling. Metabolomics data of the 6 individuals assigned to the KD who underwent metabolomic profiling was kept for further analysis. Further information, including exclusion criteria can be found at ClinicalTrials.gov (NCT03285152). Dietary composition can be found in **Supplementary Table 2**.

#### NHSII

Study participants were women from the Nurses’ Health Study II (NHSII), a long-term prospective cohort initiated 1989, enrolling nurses aged 25-42 years at baseline. Participants were selected as part of a nested case–control study on breast cancer. The NHSII group consisted of 2,117 women who provided samples between 1996 and 1999, all of whom had metabolomic profiling performed. Cases were women diagnosed with breast cancer after blood collection and before 2011. Control participants were matched to cases by age (±1 year), date of blood draw (±1 month), fasting status (≥8 h since a meal vs. <8 h or unknown), as well as menopausal status and hormone use (premenopausal, postmenopausal hormone user, postmenopausal non-user, unknown) at blood draw. Breast cancer cases were identified by self-report and confirmed by medical record review. ER status was determined via a central review of breast tissue microarrays or by pathology reports, if tissue was unavailable. Risk-factor and covariate information was collected in both cohorts via biennial questionnaires.

Covariates included established breast cancer risk factors as potential confounders The following covariates were selected at the time of blood draw from corresponding questionnaires or prior reports for non-time-varying covariates: age at menarche, age at first birth and parity combined (nulliparous, 1–2 kids & <25 y, 1–2 kids & 25 + y, 3+ kids & <25 y, 3+kids & 25 + y), history of benign breast disease (yes/no), family history of breast cancer (yes/no), BMI at age 18 (kg/m^2^), alcohol consumption at blood draw (g/day), alternative healthy eating index at blood draw (AHEI)^39^, physical activity level at blood draw (metabolic equivalent of task [MET]-h/week) and weight change since age 18 at blood draw (kg). In the case of missing values of weight change since age 18, physical activity, alcohol intake and AHEI, values were imputed with median values. Subjects missing menarche and nulliparity information were removed (N = 3 samples removed).

The study protocol was approved by the institutional review boards of the Brigham and Women’s Hospital and Harvard T.H. Chan School of Public Health, and those of participating registries as required.

### 2.2 Metabolomic Profiling

Metabolites for all three cohorts described above were profiled by Dr. Clary Clish’s lab at the Broad Institute of MIT and Harvard (Cambridge, MA) as previously described^40^. Briefly, hydrophilic interaction liquid chromatographic analyses of water-soluble metabolites in positive (HILIC-pos) and negative (HILIC-neg) ionization modes were conducted along with positive ion mode analyses of polar and non-polar plasma lipids (C8-pos) and negative ion mode analyses of free fatty acids and bile acids (C18-neg) using a liquid chromatography-tandem mass spectrometry (LC-MS/MS) system. For each method, metabolite identities were confirmed using authentic reference standards or reference samples. In the OSU and MSK cohorts, each subject’s longitudinal samples were shuffled and then run sequentially. In the NHSII cohort, matched case-control pairs were distributed randomly within batches. Pooled reference samples were included every 20 samples and blinded quality control samples were also randomly distributed. Each sample measurement was standardized by first dividing it by the value of the closest pooled reference sample in the sequence of samples. The resulting ratio was then multiplied by the median of all reference values for that metabolite.

### 2.3 Data pre-processing

For all datasets (OSU, MSK, and NHSII) preprocessing steps were performed using the maplet R package^41^ as follows: First, metabolites whose QC measurements had a coefficient of variation > 25% were filtered out. Next, metabolites with >25% missing values were filtered out followed by samples with >25% missingness (N = 2 samples removed). Probabilistic quotient normalization was applied to correct for sample-wise variation^42^ followed by log2 transformation. All outliers with abundance levels above *q = abs(qnorm[0.0125/n])*, with *n* representing the number of samples, were set to missing. Missing values were then imputed using a k-nearest neighbors imputation method^43^ and data was scaled^44^. To avoid repeated measurements of the same metabolite from different liquid chromatography methods, of the HMDB IDs that occurred more than once, the one with lowest QC coefficient of variation was kept.

To control for potential confounding, BMI and sex were regressed out of the dietary intervention cohorts by fitting a linear model, and the residuals were extracted for further analysis. These residuals represent the variation in the data that was independent of the regressed variables and were subsequently used in downstream analyses. Metabolites that significantly differed between KD and LFD subjects in week 0 (pre-intervention) of the dietary interventions were filtered out (t-test, Bonferroni adjusted p-value < 0.05)^45^. Finally, the ComBat method from the sva package in R^46^ was used to adjust for batch effects between the OSU and MSK cohorts in order to remove potential differences between metabolite profiles based on study.

The NHSII cohort had blood samples that were subject to delayed processing; metabolites impacted by delayed sample processing were removed (ICC and Spearman rho <0.75 comparing immediately vs. within 24–48-h post collection). Batch run was regressed out from the NHSII cohort and residuals were kept using the same method described above. After pre-processing, 2,112 subjects from the NHSII cohort were used for downstream analysis.

### 2.4 Statistical Analysis

#### Derivation of diet scores

For each diet group (KD and LFD), a metabolome-wide association study (MWAS) was performed using a paired t-test on metabolite concentrations at the first week (pre-intervention) vs. the final week of dietary intervention. The log-fold changes of significantly impacted metabolites (FDR-adjusted p < 0.05) were collected. A metabolomics score was then derived for both KD and LFD as the sum of each metabolite’s abundance multiplied by its respective diet-induced fold-change. Notably, since the metabolomics data was scaled, log fold changes represent a standardized measure of effect size in this context.

#### Association of diet scores with breast cancer risk

Quartiles of metabolomics scores were calculated in the control populations of the NHSII cohort for KD and LFD. Odds ratios (OR) and 95% confidence intervals (CIs) were calculated using conditional logistic regressions, adjusting for matching factors and breast cancer risk factors described in **Methods 2.1**. To examine the mediating effects of weight change on the association between diet score and breast cancer incidence, two models were designed—one including BMI at age 18 and weight change since age 18 as covariates, and one excluding them. Tests for trend were conducted using the Wald test^47^, where the medians of the quartiles were modeled continuously. To examine the impact of ER status, menopausal status at blood draw, menopausal status at cancer diagnosis, and tumor invasiveness on the relationship between breast cancer risk and diet scores, unconditional logistic regression models were run with cases in the specific stratification group and all controls. Cochran’s Q test was used to assess heterogeneity between stratified models.

#### Score assessment

Given the substantial differences in context between the controlled dietary-intervention trials (OSU, MSK) and the observational NHSII cohort, we assessed how well the KD and LFD metabolomics scores derived from the controlled feeding studies captured similar metabolic states in an observational cohort. To this end, we defined an *observed effect* value for each metabolite that reflects the relationship between each metabolite and the full diet score in NHSII. This observed effect value was calculated as the Pearson correlation coefficient between the metabolite’s abundance and the diet score across subjects. A high absolute observed effect indicates that a metabolite’s abundance contributes strongly to the diet score, and thus the metabolite strongly contributes to the score’s stratification of subjects in NHSII.

The observed effect of each metabolite was compared to its corresponding metabolite-specific fold-change derived from the dietary intervention trials, allowing us to compare the expected contribution of a metabolite to the diet score (its fold-change multiplier) to its actual contribution to the diet score (its observed effect). We then correlated these observed effect values with the fold-changes across all metabolites to quantify the degree of divergence between the feeding trials (OSU, MSK) and the observational study (NHSII).

#### Association of individual metabolites with breast cancer risk

To assess how individual metabolites within the dietary metabolomic scores contributed to the association between the diet score and breast cancer risk, each metabolite was individually included in a conditional logistic regression model with breast cancer incidence as the outcome. The multivariable model outlined above adjusting for matching factors, established breast cancer risk factors, and BMI related parameters was run on each metabolite.

All analyses were performed with R 4.2.0.

## 3 Results

### 3.1 Metabolomic composition of dietary scores

Of 215 metabolites measured with known metabolite identities, widespread changes were observed for both the KD and LFD, with 114 and 121 significant diet-associated metabolites, respectively (paired t-test, FDR-adjusted p-value < 0.05, **Supplementary Data 1**). There was a large overlap of these changes across the two diets, with 68 metabolites significantly affected in the same direction by both diets and 6 in divergent directions.

As expected, the metabolites with the greatest positive fold-changes on the KD were the ketone body beta-hydroxybutyrate and its hydroxybutyrylcarnitine counterpart (**Figure 2a**, C4-OH carnitine). These two metabolites were both also significantly increased in the LFD (**Figure 2a**), suggesting ketosis as a general feature of hypocaloric diets. Both diets also led to similar patterns of changes across various lipid groups, namely cholesterol esters (CEs), phosphatidylcholines (PCs), plasmalogens, and sphingomyelins (SMs, **Figure 2b**). A universal pattern appeared across these lipid species for both diets: shorter chain lipids with fewer double bonds (higher degree of saturation) decreased in abundance, while longer chain lipids with more double bonds (lower degree of saturation) increased. This pattern is further apparent when examining changes in triglycerides for both diets (**Figure 2b**).

**Figure 2.**
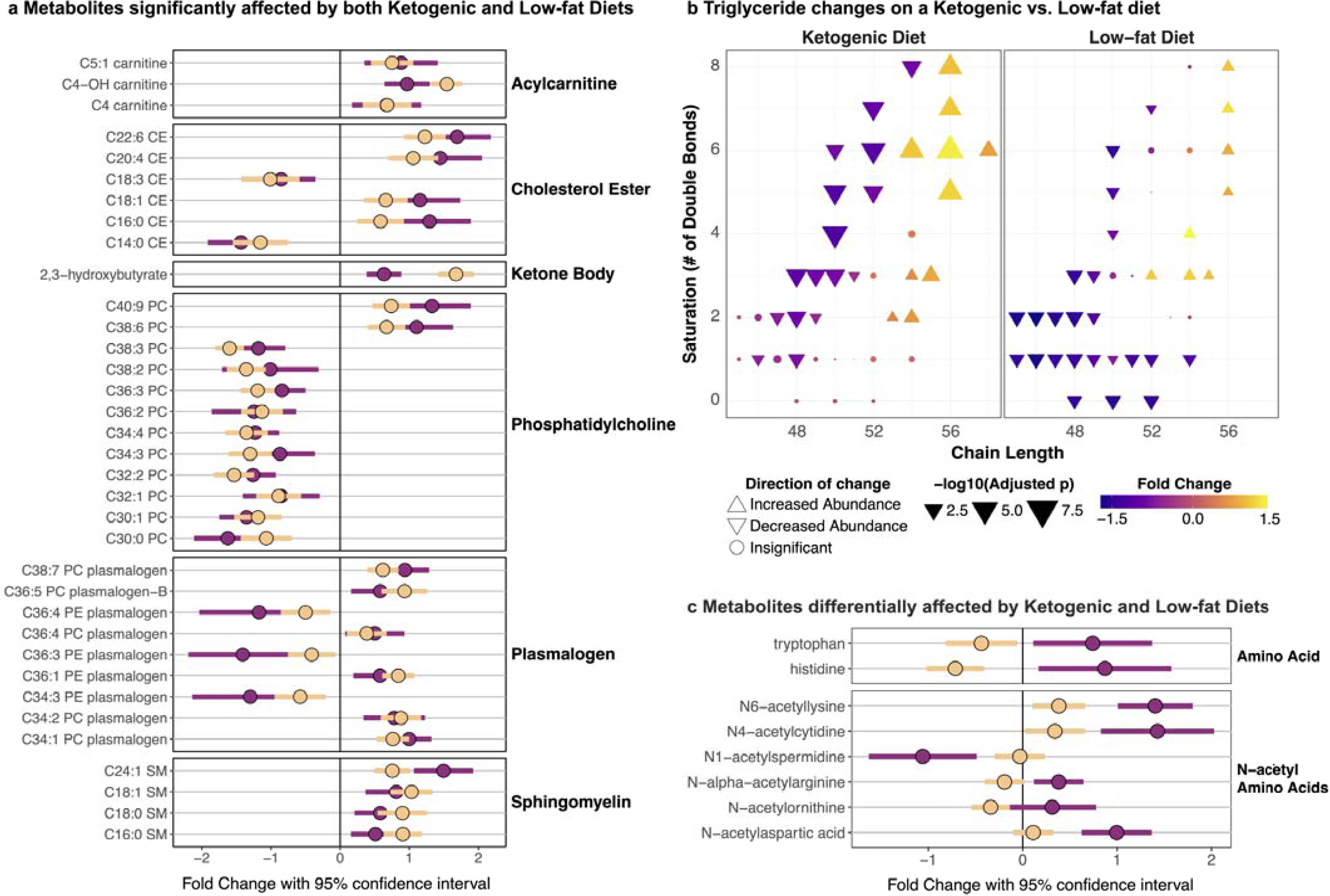
Impact of hypocaloric diets on metabolites. **a** Fold changes and 95% confidence intervals of metabolites due to the keto diet (yellow) and the low-fat diet (purple). Metabolites are organized by their molecular class. **b** Triglyceride changes on the ketogenic vs. low-fat diet, with color indicating fold-change, size indicating significance level, and shape indicating direction of change. Triglyceride chain length is represented on the x-axis, while the number of double bonds (degree of saturation) is on the y-axis. There is a clear pattern in both diets that shows an increase in the abundance of long-chain, unsaturated triglycerides while a decrease in the abundance of shorter chain, more saturated (fewer double bonds) triglycerides. **c** N-acetylated amino acids were more strongly affected by low-fat diet than ketogenic diet, while tryptophan and histidine changed in different directions between diets

Despite the similarities in metabolite changes across the two diets, there were divergent patterns as well. There was a substantially larger increase of polyunsaturated, long-chain triglycerides in the KD as compared to the LFD, which showed a greater decrease of abundance among short chain triglycerides with few double bonds (**Figure 2b**). In addition, the low-fat diet had a greater impact on amino acids, increasing tryptophan and histidine which decreased on the KD, and increasing most N-acetylated amino acids (no effect in KD, **Figure 2c**).

The diet scores, computed using fold-changes of the significantly changed metabolites, demonstrated strong separation of subjects between week 0 (pre-intervention) and weeks 1–5 (during intervention), as expected, with an AUC of 0.99 for KD and 0.93 for LFD (**Supplementary** Figure 1).

### 3.2 NHSII Descriptive Characteristics

The analysis included 1,058 incident breast cancer cases and 1,054 controls from the NHSII cohort. Descriptive characteristics are included in **Table 1** for NHSII controls according to the quartiles of participant ketogenic diet scores; the table for NHSII characteristics stratified by LFD scores can be found in **Supplementary Table 3**. The average age of participants at blood collection was 44.7 years (SD = 4.5). 72% of participants were fasting at blood draw, and 77% participants were premenopausal at blood draw (42% premenopausal at diagnosis). Participants in higher KD and LFD score quartiles had lower rates of breast cancer family history and less weight gain since age 18.

**Table 1.** Descriptive characteristics of Nurses’ Health Study II participants, tabulated by Ketogenic Diet metabolomics score quartiles among controls (N = 1054)

| Characteristic | Quartile1<br>N = 264<br>( < -20.81) | Quartile 2<br>N = 263<br>( ≥ -20.81 to < -2.59) | Quartile 3<br>N = 263<br>( ≥ -2.59 to < 17.41) | Quartile 4<br>N = 264<br>( ≥ 17.41) | P-value<sup>1</sup> |
| --- | --- | --- | --- | --- | --- |
| **Age at blood draw (Years), mean (SD)** | 45.9 (4.2) | 44.7 (4.6) | 44.7 (4.2) | 44.1 (4.5) | 0.509 |
| **Fasting at blood draw (>8 hours), N (%)** | 192 (73%) | 189 (72%) | 204 (78%) | 205 (78%) | 0.26 |
| **Menopausal status and PMH use at blood draw, N (%)** |  |  |  |  | 0.042 |
| Postmenopausal – no hormone use | 7 (2.7%) | 7 (2.7%) | 7 (2.7%) | 2 (0.8%) | - |
| Postmenopausal – yes hormone use | 38 (14%) | 22 (8.4%) | 25 (9.5%) | 30 (11%) | - |
| Premenopausal | 184 (70%) | 204 (78%) | 206 (78%) | 215 (81%) | - |
| Unknown | 35 (13%) | 30 (11%) | 25 (9.5%) | 17 (6.4%) | - |
| **Age at Menarche (Years), mean (SD)** | 12.4 (1.5) | 12.3 (1.3) | 12.5 (1.4) | 12.7 (1.5) | 0.032 |
| **Nulliparous, N (%)** | 47 (18%) | 40 (15%) | 52 (20%) | 49 (19%) | 0.571 |
| **Parity, mean (SD)** | 2.3 (1) | 2.5 (1.1) | 2.3 (1) | 2.3 (0.9) | 0.581 |
| **History of benign breast disease, N (%)** | 44 (17%) | 47 (18%) | 35 (13%) | 38 (14%) | 0.457 |
| **Family history of breast cancer, N (%)** | 39 (15%) | 30 (11%) | 27 (10%) | 18 (6.8%) | 0.032 |
| **BMI at age 18 (kg/m<sup>2</sup>), mean (SD)** | 21.5 (3.2) | 21 (2.9) | 21 (3) | 21.2 (3.2) | 0.759 |
| **Weight change since age 18 (kg), mean (SD)** | 17 (15) | 13 (11) | 11 (12) | 10 (11) | 0.0003 |
| **Activity levels at blood draw (MET-hours/week), mean (SD)** | 18 (18) | 17 (20) | 19 (30) | 19 (20) | 0.867 |
| **Current Smoker, N (%)** | 21 (8%) | 15 (5.7%) | 22 (8.4%) | 16 (6.1%) | 0.541 |
| **Alcohol Consumption at blood draw (g/day), mean (SD)** | 3.1 (5.8) | 2.9 (5.5) | 4 (7.4) | 3.3 (5.4) | 0.534 |

<sup>1</sup> Nominal p-values after testing for differences between quartiles. For continuous variables (age at blood draw, age at menarche, parity, BMI at age 18, weight change since age 18, activity levels at blood draw, and alcohol consumption at blood draw), a Kruskal-Wallis test was used. For all other variables, a chi-squared test was used.

### 3.3 Association of diet scores with breast cancer risk

We observed statistically significant positive associations between both KD and LFD diet scores and overall breast cancer risk in the NHSII cohort (**Tables 2 and 3**). Women in the highest quartile of KD score (i.e., with a profile similar to post-diet intervention) had a 37% increased risk of breast cancer (OR=1.37, 95% CI 1.06-1.77; p-trend=0.021) while women in the highest quartile of LFD score had a 32% increased risk (OR=1.32, 95% CI 1.02-1.7; p-trend=0.01) compared to their respective lowest quartiles. This risk increase was even higher in invasive breast cancer cases (44% and 47% increased risk in highest quartiles of KD and LFD, respectively) and ER+ cases (53% and 50% increased risk, respectively), though there was no significant difference in association between ER status (positive vs. negative) or tumor invasiveness (invasive vs. in situ) (Cochran’s Q-test, p > 0.05, **Tables 2 and 3**).

**Table 2.** Odds ratios and 95% Confidence Intervals for quartiles of ketogenic diet metabolomics scores and breast cancer risk in Nurses’ Health Study II (1,058 cases/1,054 controls)

| **Ketogenic Diet Metabolomics Score** | **Categorical OR (95% CI)** |  |  |  |  | **Continuous OR (95% CI)** |  |  |
| --- | --- | --- | --- | --- | --- | --- | --- | --- |
| **Overall breast cancer (1,058 cases)** | Q1 | Q2 | Q3 | Q4 | P-trend<sup>4</sup> | Per SD increase | P-value | P-het<sup>5</sup> |
| Multivariable adjusted<sup>1</sup> | 1.0 (ref) | 1.28 (0.98, 1.66) | 1.31 (1.02, 1.68) | 1.37 (1.06, 1.77) | 0.02 | 1.1 (1.01, 1.21) | 0.032 | - |
| Multivariable adjusted + BMI<sup>2</sup> | 1.0 (ref) | 1.25 (0.96, 1.63) | 1.27 (0.98, 1.64) | 1.33 (1.02, 1.73) | 0.05 | 1.09 (1, 1.2) | 0.060 | - |
| **ER-positive breast cancer (585 cases)** |  |  |  |  |  |  |  |  |
| Multivariable adjusted<sup>3</sup> | 1.0 (ref) | 1.37 (1.01, 1.86) | 1.37 (1.01, 1.86) | 1.53 (1.13, 2.07) | 0.01 | 1.15 (1.04, 1.27) | 0.011 | 0.29 |
| Multivariable adjusted + BMI<sup>2,3</sup> | 1.0 (ref) | 1.34 (0.99, 1.82) | 1.34 (0.98, 1.83) | 1.5 (1.1, 2.04) | 0.02 | 1.14 (1.03, 1.26) | 0.016 | 0.29 |
| **ER-negative breast cancer (153 cases)** |  |  |  |  |  |  |  |  |
| Multivariable adjusted<sup>3</sup> | 1.0 (ref) | 1 (0.61, 1.63) | 0.98 (0.59, 1.61) | 1.16 (0.72, 1.89) | 0.55 | 1.03 (0.86, 1.23) | 0.724 | - |
| Multivariable adjusted + BMI<sup>2,3</sup> | 1.0 (ref) | 0.96 (0.58, 1.57) | 0.92 (0.56, 1.54) | 1.1 (0.67, 1.8) | 0.70 | 1.02 (0.86, 1.22) | 0.863 | - |
| **Premenopausal at blood draw (819 cases)** |  |  |  |  |  |  |  |  |
| Multivariable adjusted<sup>3</sup> | 1.0 (ref) | 1.15 (0.85, 1.54) | 1.32 (0.99, 1.77) | 1.31 (0.98, 1.74) | 0.05 | 1.11 (1, 1.22) | 0.044 | 0.79 |
| Multivariable adjusted + BMI<sup>2,3</sup> | 1.0 (ref) | 1.11 (0.82, 1.49) | 1.28 (0.95, 1.71) | 1.26 (0.94, 1.69) | 0.09 | 1.09 (0.99, 1.21) | 0.071 | 0.79 |
| **Postmenopausal at blood draw (239 cases)** |  |  |  |  |  |  |  |  |
| Multivariable adjusted<sup>3</sup> | 1.0 (ref) | 1.49 (0.9, 2.47) | 1.36 (0.8, 2.31) | 1.65 (0.97, 2.83) | 0.09 | 1.14 (0.94, 1.39) | 0.191 | - |
| Multivariable adjusted + BMI<sup>2,3</sup> | 1.0 (ref) | 1.44 (0.86, 2.4) | 1.35 (0.79, 2.31) | 1.59 (0.92, 2.74) | 0.12 | 1.13 (0.93, 1.37) | 0.229 | - |
| **Premenopausal at diagnosis (446 cases)** |  |  |  |  |  |  |  |  |
| Multivariable adjusted<sup>3</sup> | 1.0 (ref) | 1.24 (0.86, 1.78) | 1.49 (1.04, 2.12) | 1.44 (1.01, 2.05) | 0.03 | 1.15 (1.02, 1.29) | 0.02 | 0.52 |
| Multivariable adjusted + BMI<sup>2,3</sup> | 1.0 (ref) | 1.19 (0.83, 1.72) | 1.43 (1, 2.05) | 1.38 (0.97, 1.98) | 0.06 | 1.14 (1.01, 1.28) | 0.032 | 0.52 |
| **Postmenopausal at diagnosis (612 cases)** |  |  |  |  |  |  |  |  |
| Multivariable adjusted<sup>3</sup> | 1.0 (ref) | 1.19 (0.89, 1.61) | 1.23 (0.91, 1.66) | 1.35 (1, 1.81) | 0.05 | 1.09 (0.99, 1.21) | 0.106 | - |
| Multivariable adjusted + BMI<sup>2,3</sup> | 1.0 (ref) | 1.15 (0.85, 1.55) | 1.19 (0.88, 1.61) | 1.31 (0.97, 1.78) | 0.08 | 1.08(0.98, 1.19) | 0.149 | - |
| **In situ breast cancer (276 cases)** |  |  |  |  |  |  |  |  |
| Multivariable adjusted<sup>3</sup> | 1.0 (ref) | 0.98 (0.65, 1.48) | 1.39 (0.94, 2.04) | 1.2 (0.8, 1.79) | 0.20 | 1.09 (0.95, 1.26) | 0.205 | 0.82 |
| Multivariable adjusted + BMI<sup>2,3</sup> | 1.0 (ref) | 0.92 (0.61, 1.4) | 1.33 (0.9, 1.97) | 1.14 (0.76, 1.71) | 0.29 | 1.08 (0.94, 1.24) | 0.283 | 0.73 |
| **Invasive breast cancer (738 cases)** |  |  |  |  |  |  |  |  |
| Multivariable adjusted<sup>3</sup> | 1.0 (ref) | 1.27 (0.96, 1.68) | 1.27 (0.96, 1.68) | 1.44 (1.09, 1.9) | 0.01 | 1.12 (1.01, 1.23) | 0.022 | - |
| Multivariable adjusted + BMI<sup>2,3</sup> | 1.0 (ref) | 1.24 (0.94, 1.64) | 1.24 (0.93, 1.64) | 1.4 (1.06, 1.86) | 0.03 | 1.12 (1.01, 1.23) | 0.037 | - |

<sup>1</sup> conditional logistic regression adjusted for smoking, alcohol intake, physical activity, age at menarche, parity and age of first birth, family history of breast cancer, and personal history of benign breast disease
<sup>2</sup> additionally adjusted for BMI at age 18 and weight change since age 18 to blood draw.
<sup>3</sup> unconditional logistic regression adjusted for variables in **1** and matching variables including age, fasting status, menopausal status at blood draw, and blood draw date
<sup>4</sup> tests for trend were conducted using the Wald test where the medians of the quartiles were modeled continuously
<sup>5</sup> tests for heterogeneity were performed using a Cochran's Q-test

**Table 3.** Odds ratios and 95% Confidence Intervals for quartiles of low-fat diet metabolomics scores and breast cancer risk in Nurses’ Health Study II (1,058 cases/1,054 controls)

| Low Fat Diet Metabolomics Score | Categorical OR (95% CI) |  |  |  |  | Continuous OR (95% CI) |  |  |
| --- | --- | --- | --- | --- | --- | --- | --- | --- |
|  | Q1 | Q2 | Q3 | Q4 | P-trend<sup>4</sup> | Per SD increase | P-value | P-het<sup>5</sup> |
| **Overall breast cancer (1,058 cases)** |  |  |  |  |  |  |  |  |
| Multivariable adjusted<sup>1</sup> | 1.0 (ref) | 1.09 (0.84, 1.42) | 1.45 (1.13, 1.87) | 1.32 (1.02, 1.7) | 0.01 | 1.09 (1, 1.2) | 0.053 | - |
| Multivariable adjusted + BMI<sup>2</sup> | 1.0 (ref) | 1.04 (0.79, 1.36) | 1.39 (1.07, 1.81) | 1.24 (0.96, 1.62) | 0.03 | 1.07 (0.98, 1.18) | 0.131 | - |
| **ER-positive breast cancer (585 cases)** |  |  |  |  |  |  |  |  |
| Multivariable adjusted<sup>3</sup> | 1.0 (ref) | 1.12 (0.82, 1.52) | 1.48 (1.09, 2) | 1.5 (1.1, 2.03) | 0.00 | 1.15 (1.02, 1.29) | 0.009 | 0.52 |
| Multivariable adjusted + BMI<sup>2,3</sup> | 1.0 (ref) | 1.09 (0.8, 1.49) | 1.46 (1.07, 1.99) | 1.45 (1.06, 1.99) | 0.01 | 1.15 (1.02, 1.29) | 0.018 | 0.44 |
| **ER-negative breast cancer (153 cases)** |  |  |  |  |  |  |  |  |
| Multivariable adjusted<sup>3</sup> | 1.0 (ref) | 1.05 (0.63, 1.75) | 1.3 (0.79, 2.14) | 1.32 (0.8, 2.19) | 0.21 | 1.07 (0.9, 1.28) | 0.421 | - |
| Multivariable adjusted + BMI<sup>2,3</sup> | 1.0 (ref) | 1 (0.59, 1.68) | 1.22 (0.73, 2.06) | 1.24 (0.73, 2.11) | 0.32 | 1.05 (0.86, 1.28) | 0.596 | - |
| **Premenopausal at blood draw (819 cases)** |  |  |  |  |  |  |  |  |
| Multivariable adjusted<sup>3</sup> | 1.0 (ref) | 1 (0.74, 1.36) | 1.41 (1.05, 1.89) | 1.32 (0.98, 1.77) | 0.02 | 1.08 (0.98, 1.19) | 0.124 | 0.33 |
| Multivariable adjusted + BMI<sup>2,3</sup> | 1.0 (ref) | 0.95 (0.7, 1.3) | 1.36 (1, 1.84) | 1.25 (0.92, 1.7) | 0.04 | 1.06 (0.96, 1.17) | 0.245 | 0.24 |
| **Postmenopausal at blood draw (239 cases)** |  |  |  |  |  |  |  |  |
| Multivariable adjusted<sup>3</sup> | 1.0 (ref) | 1.39 (0.85, 2.26) | 1.59 (0.94, 2.69) | 1.49 (0.86, 2.58) | 0.09 | 1.21 (0.99, 1.47) | 0.051 | - |
| Multivariable adjusted + BMI<sup>2,3</sup> | 1.0 (ref) | 1.34 (0.81, 2.2) | 1.54 (0.9, 2.65) | 1.44 (0.82, 2.52) | 0.13 | 1.21 (0.99, 1.47) | 0.067 | - |
| **Premenopausal at diagnosis (446 cases)** |  |  |  |  |  |  |  |  |
| Multivariable adjusted<sup>3</sup> | 1.0 (ref) | 1.04 (0.71, 1.52) | 1.63 (1.14, 2.34) | 1.43 (1, 2.06) | 0.01 | 1.14 (1.01, 1.28) | 0.037 | 0.52 |
| Multivariable adjusted + BMI<sup>2,3</sup> | 1.0 (ref) | 0.99 (0.67, 1.45) | 1.56 (1.09, 2.26) | 1.36 (0.93, 1.97) | 0.03 | 1.13 (0.98, 1.29) | 0.077 | 0.59 |
| **Postmenopausal at diagnosis (612 cases)** |  |  |  |  |  |  |  |  |
| Multivariable adjusted<sup>3</sup> | 1.0 (ref) | 1.1 (0.82, 1.47) | 1.38 (1.02, 1.85) | 1.34 (0.99, 1.81) | 0.03 | 1.08 (0.98, 1.19) | 0.136 | - |
| Multivariable adjusted + BMI<sup>2,3</sup> | 1.0 (ref) | 1.05 (0.77, 1.42) | 1.34 (0.99, 1.820) | 1.29 (0.94, 1.76) | 0.05 | 1.07 (0.95, 1.21) | 0.229 | - |
| **In situ breast cancer (276 cases)** |  |  |  |  |  |  |  |  |
| Multivariable adjusted<sup>3</sup> | 1.0 (ref) | 0.91 (0.6, 1.39) | 1.47 (0.99, 2.18) | 1.09 (0.72, 1.64) | 0.29 | 1.02 (0.89, 1.17) | 0.748 | 0.2 |
| Multivariable adjusted + BMI<sup>2,3</sup> | 1.0 (ref) | 0.85 (0.56, 1.3) | 1.37 (0.91, 2.05) | 1.02 (0.67, 1.56) | 0.45 | 1 (0.85, 1.17) | 0.999 | 0.2 |
| **Invasive breast cancer (738 cases)** |  |  |  |  |  |  |  |  |
| Multivariable adjusted<sup>3</sup> | 1.0 (ref) | 1.1 (0.83, 1.46) | 1.42 (1.08, 1.88) | 1.47 (1.11, 1.94) | 0.00 | 1.14 (1.03, 1.26) | 0.012 | - |
| Multivariable adjusted + BMI<sup>2,3</sup> | 1.0 (ref) | 1.06 (0.8, 1.42) | 1.4 (1.05, 1.87) | 1.41 (1.05, 1.89) | 0.01 | 1.13 (1.02, 1.24) | 0.027 | - |

<sup>1</sup> conditional logistic regression adjusted for smoking, alcohol intake, physical activity, age at menarche, parity and age of first birth, family history of breast cancer, and personal history of benign breast disease
<sup>2</sup> additionally adjusted for BMI at age 18 and weight change since age 18 to blood draw.
<sup>3</sup> unconditional logistic regression adjusted for variables in 1 and matching variables including age, fasting status, menopausal status at blood draw, and blood draw date
<sup>4</sup> tests for trend were conducted using the Wald test where the medians of the quartiles were modeled continuously
<sup>5</sup> tests for heterogeneity were performed using a Cochran's Q-test

The association between both metabolomics diet scores and breast cancer risk did not significantly differ by menopausal status at blood draw or cancer diagnosis (Cochran’s Q-test, p > 0.05, **Tables 2 and 3**). However, the associations between metabolomics scores were attenuated slightly when we further adjusted for BMI at age 18 and weight change since age 18 (KD OR=1.33, 95% CI 1.02-1.73and LFD OR=1.24, 95% CI 0.96-1.62, **Tables 2 and 3**). To assess the impact of fasting on the association between scores and BC risk, sensitivity analysis using only fasted samples was performed; the association between metabolomics scores and breast cancer remained unchanged (KD OR = 1.37, 95% CI 1.01-1.91, LFD OR = 1.39, 95% CI 1-1.89, **Supplementary Table 4 and 5**).

### 3.4 Assessment of metabolomics score in NHSII

Given the substantial differences in context between the controlled dietary-intervention trials and the NHSII cohort, we sought to assess how well the dietary metabolomics scores captured similar metabolic states in the observational cohort. To this end, we determined how much each metabolite contributed to the diet score in the NHSII setting by generating observed effect values. The observed effect of a metabolite was defined as the Pearson correlation coefficient between a metabolite’s abundance and the diet score across subjects. These observed effects in NHSII closely matched their respective metabolite fold changes calculated in the controlled trials (KD r = 0.742, 95% CI 0.647–0.815; LFD r = 0.821, 95% CI 0.753–0.872, **Figure 3a,b**). This agreement indicates that the majority of metabolites in the diet score were stratifying NHSII subjects in the same way, and by the same metabolic states, as in the controlled diet trials.

**Figure 3.**
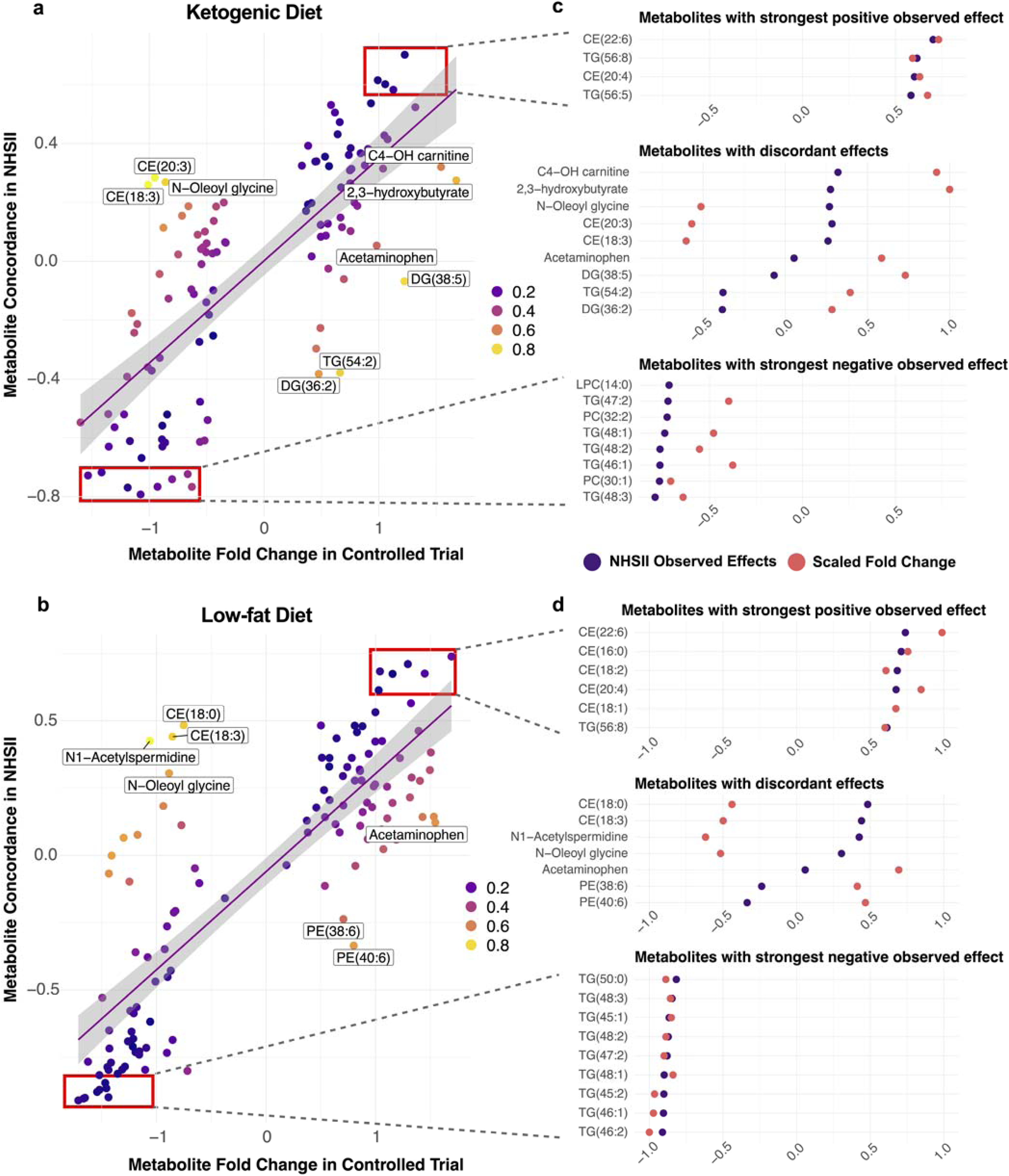
Comparison between metabolite observed effect values in NHSII observational cohort and metabolite fold change from controlled trials. **a** Scatter plot of metabolite fold change (x-axis) and metabolite observed effect on diet scores in NHSII (y-axis) for KD and **b** LFD. Color represents absolute difference between the observed effect and the scaled fold-change, and select metabolites that fall far from the line are labelled **c** Metabolites with the strongest positive and negative observed effecton NHS diet scores (within red boxes of scatterplots), as well as metabolites with discordant effects between study settings in the KD diet scores and **d** LFD diet scores. Purple circles represent NHSII observed effect values, orange circles represent scaled fold changes from the controlled trials. When an observed effect value and fold change value are close, that indicates the metabolite contributed to the diet score to the same degree in the two settings.

The metabolites that most strongly contributed to the stratification of NHSII subjects for both diets were those with the largest absolute observed effect (**Figure 3c,d**). Of the highest positive observed effect values in the NHSII cohort, cholesterol esters were prominently represented across both diets, reflecting their large contribution to the stratification of NHSII subjects by diet score. Notably, C22:6 CE showed the strongest positive association with both scores. Long-chain, polyunsaturated triglycerides also exhibited strong positive observed effects, and thus contribution to both diet scores, while plasmalogens strongly contributed to only the KD score. Conversely, short-chain, less saturated triglycerides had the strongest negative observed effect on both diet scores, with a markedly higher representation in the LFD compared to the KD. In contrast, the strongest negative observed effects in the KD score included numerous phosphatidylcholines (**Figure 3c**).

Several metabolites showed a drastic discordance between their effect in NHSII and their fold-change in the diet scores (**Figure 3c,d**). This indicates that these metabolites did not contribute to the diet scores in NHSII to the same extent as the controlled trials, revealing how the different contexts affected individual metabolites within the diet scores. Interestingly, the ketone body beta-hydroxybutyrate and its carnitine counterpart—both of which had the highest fold-changes and, consequently, the largest multipliers in the KD score—showed only modest contributions to the KD scores in the NHSII cohort. This finding suggests the stratification of NHSII participants by diet score is not as largely driven by ketone body production as the controlled trial subjects.

This pattern was also true of acetaminophen across both diets. While the multiplier of acetaminophen was positive in the controlled trials, its contribution to the diet scores dropped to near zero in the NHSII cohort. This indicates that many subjects in the dietary intervention setting may have been taking the pain reliever as the study progressed, but acetaminophen does not reflect the metabolic state in the observational population.

### 3.5 Individual Metabolite Contributions to Risk Association

To determine how individual metabolites within the dietary metabolomic scores specifically contributed to breast cancer risk, each metabolite was analyzed separately in a multivariable conditional logistic regression model with breast cancer incidence as the outcome (see **Methods 2.4**). The metabolites with the strongest absolute associations to breast cancer risk that were significantly impacted by both diets included hydroxybutyrylcarnitine (C4-OH carnitine), N-Oleoyl glycine, CE(22:6), and some short-chain triglycerides with high saturation (**Figure 4a,b**). The regression coefficients for N-Oleoyl glycine differed in direction from the dietary intervention fold change. KD also impacted LPC(20:1), a long chain plasmalogen (PE(P-40:6)), and LPE(22:0), while LFD had a moderate effect on Piperidine and Cotinine which were negatively associated with BC risk.

**Figure 4.**
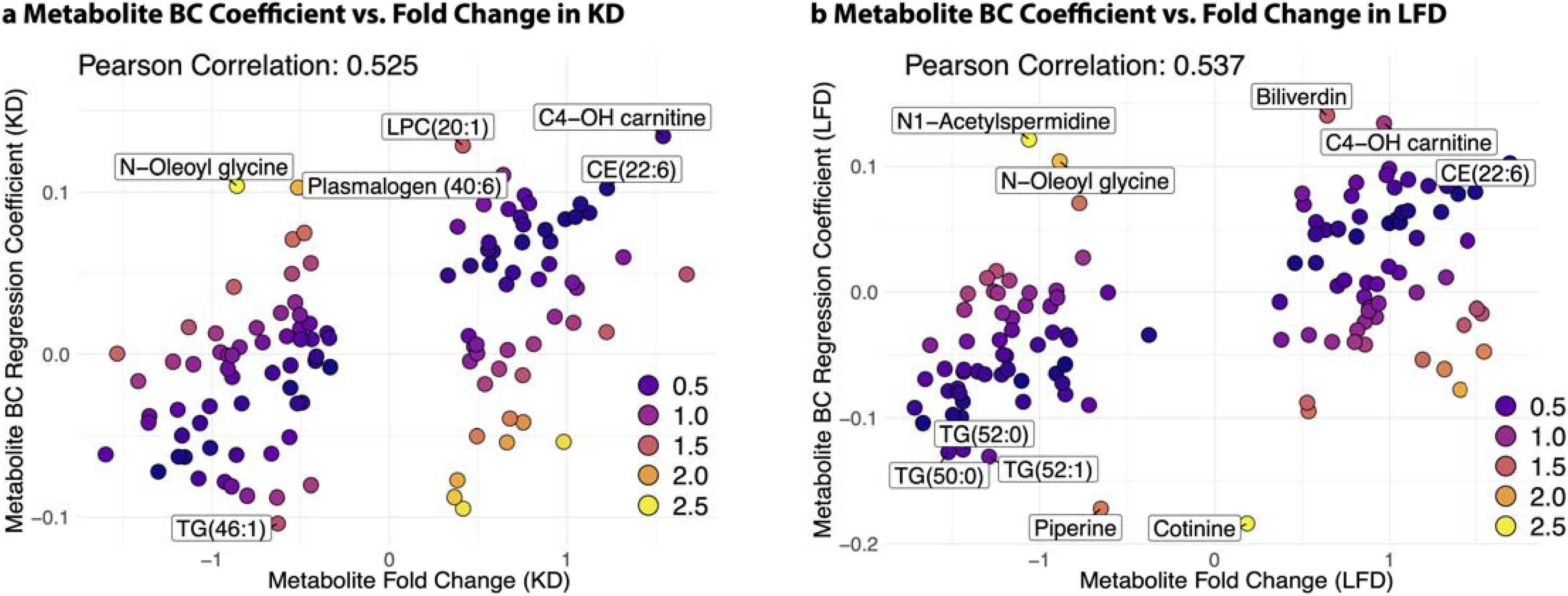
Comparison of metabolites in diet scores and their BC risk regression coefficients. **a** Metabolite fold changes on the ketogenic diet vs BC regression coefficient. **b** Metabolite fold changes on low-fat diet vs BC regression coefficient. Both comparisons show positive correlations, indicating metabolites systematically contribute to their respective diet scores in the same pattern that they contribute to breast cancer risk in the NHSII cohort. Point colors represent absolute difference between the scaled fold-change and the scaled regression coefficient.

To examine if the metabolomics diet score’s association with breast cancer risk was driven by a few metabolites or due to a global pattern, we examined the relationship between all metabolite fold-changes with their regression coefficients. Across both dietary patterns, there was a moderate positive correlation between the fold-changes of each metabolite and their corresponding regression coefficients. For the KD score, the Pearson correlation between metabolite fold-changes and regression coefficients was 0.52 (95% CI 0.38–0.65; **Figure 4a**). Similarly, for the LFD score, there was a positive correlation between metabolite fold-changes and regression coefficients (Pearson correlation of 0.54, 95% CI 0.40–0.65; **Figure 4b**). These positive correlations show that the metabolic states of hypocaloric diet systematically increase metabolites positively associated with breast cancer risk while decreasing metabolites negatively associated with risk.

## 4 Discussion

We generated metabolomic scores from two controlled dietary intervention trials, both utilizing hypocaloric diets: one ketogenic (KD) and one low-fat (LFD). Our primary goal was to gain insight into the relationship between the metabolic state induced by these hypocaloric diets and breast cancer risk in a predominantly premenopausal cohort. Using a large, nested case-control study, we found a significant positive association between both diet scores and breast cancer risk; that is, metabolic states that appeared similar to a post-diet intervention state coincided with increased breast cancer risk. Women in the highest quartile for KD and LFD scores experienced a 37% and 32% increased breast cancer risk, respectively, compared to those in the lowest quartile. These associations were robust, with little attenuation after adjusting for confounders such as BMI at age 18 and weight change since age 18.

### 4.1 Metabolomic Signatures of KD and LFD

We found broad similarities in the metabolic response to hypocaloric diets, despite large differences in macronutrient composition of each diet. The KDs were largely composed of fat (>70% fat by kcal), while the LFD was largely composed of carbohydrates (<30% fat by kcal). Nevertheless, our longitudinal differential analysis revealed that both the KD and LFD produced similar metabolomic signatures. This finding suggests that the primary driver of the observed metabolomic changes may not be the macronutrient composition of the diets but rather the host response to the caloric restriction and resultant weight loss. As both diets had a similar association with breast cancer risk, this conclusion is consistent with prior studies showing no association between dietary fat intake and breast cancer incidence^48,49^.

Among the shared metabolic changes, the relative amounts of long-chain, polyunsaturated TGs increased in both diets, whereas short-chain, saturated TG levels decreased (more prominently in the LFD). Both diets also showed an overall increase in cholesterol esters (CEs). Both these particular patterns have been previously identified in the blood metabolome following acute starvation^50^ and may be indicative of acyl-chain remodeling from TGs to a more metabolically active pool of CEs.

While the two diets shared several common metabolic effects, key differences emerged in their TG profiles. Long-chain, polyunsaturated TGs increased more prominently in KD whereas short-chain, saturated TGs decreased to a greater extent in LFD. This divergence between diets is consistent with the dietary fat composition and metabolic demands unique to each diet. The KD’s high intake of longer-chain polyunsaturated fatty acids likely helps preserve and replenish the dietary-derived pool of long-chain, polyunsaturated TGs. Conversely, the LFD, with an overall reduced fat intake and reliance on endogenous fat stores under caloric restriction, likely promotes the utilization of both long and short-chain TGs to meet energy demands, ultimately unable to replenish long chain TGs.

### 4.2 Breast Cancer Risk and Hypocaloric Metabolic States

CEs showed a strong correlation with the diet scores in the NHSII cohort and strong positive metabolite-specific coefficients when regressed on BC incidence, indicating their increased concentration within NHSII contributed to the positive association with BC risk. Biologically, CEs serve as a critical source of cholesterol for steroidogenesis^51^. This connection is particularly relevant to estrogen receptor positive breast cancer risk, as enhanced cholesterol availability may support the biosynthesis of estrogen. Our results support this hypothesis as the diet score conferred greater breast cancer risk for NHSII ER+ cases as compared to overall cases for both KD and LFD (53% increased risk and 50% increased risk, respectively, **Tables 2 and 3**).

The widespread depletion of short-chain triglycerides in both diets are evidence that hypocaloric dietary interventions rewire systemic metabolism to promote lipolysis and release of stored lipids from TG and other phospholipids^52^. The byproducts of triglyceride lipolysis, including free fatty acids and glycerol, may serve as a link between dietary metabolomic signatures and breast cancer risk, as they have consistently been associated with metabolic rewiring in cancer cells^53,54^. Recent findings highlight a metabolic symbiosis between tumor-surrounding adipocytes and breast cancer cells, where adipocyte-derived lipids fuel tumor progression through lipolysis and fatty acid oxidation^55^. This lipid-driven metabolic remodeling enhances cancer cell invasiveness, underscoring the potential role of diet-induced lipolysis in shaping the tumor microenvironment and influencing disease development and progression.

### 4.3 Bridging study contexts with metabolomics data

This study highlights the potential of metabolomics-based dietary signatures as a novel tool for nutritional epidemiology. Unlike traditional dietary assessments, such as food diaries and recalls, which are subject to recall bias and measurement error, metabolomic profiles offer an objective and integrative measure of dietary exposure. A key innovation of this study was the use of metabolomic scores to bridge data from two fundamentally different study designs: controlled dietary intervention trials and a large, nested case-control study. This approach enabled us to transfer metabolic insights from tightly controlled experiments to a real-world epidemiological cohort, thereby combining the strengths of both methodologies.

Importantly, our findings demonstrate that diet-derived metabolomic scores effectively capture biologically meaningful metabolite associations with breast cancer risk. By comparing the fold-changes of metabolites in the controlled trial scores to their observed effect in the NHSII cohort (**Figures 2a,b**), we showed that the dietary scores largely retained their designed function of representing specific metabolic states, even in the context of an observational cohort where diet-associated metabolite variation is inherently less controlled. This observation indicates that the metabolic state of the dietary interventions is being captured by the score, and its subsequent stratification of the NHSII population is driving the association with BC risk.

The comparison of metabolite fold-changes in the controlled trials to their observed effect in the NHSII cohort also showed instances where metabolite changes in the controlled setting did not contribute to NHSII participant stratification. For example, beta-hydroxybutrate and its carnitine counterpart, while drastically impacted by the ketogenic diet, were not highly concordant in the NHSII setting (**Figures 2c,d**), suggesting the stratification of NHSII subjects was not driven by ketone body production as much as in the controlled setting. In addition, the finding that the pain killer acetaminophen, which had a positive fold-change in the controlled setting but very low effect on the diet scores in the NHSII cohort, shows that the translation of our diet score in the observational setting is robust to uncontrolled confounding metabolites within the controlled setting.

Finally, we demonstrated that the metabolite fold-changes from both diets had a strong positive correlation with the metabolites coefficients when regressed on breast cancer incidence. These findings indicate that metabolites that significantly increased in both dietary interventions also tend to be positively associated with breast cancer incidence, and vice versa. These results confirm that the metabolic state induced by both the KD and LFD systematically elevates detrimental metabolites and reduces protective metabolites with respect to breast cancer risk.

### 4.4 Limitations

This study underscores the utility of metabolomics-based scores in linking dietary patterns to disease risk; however, applying a metabolomic score derived from individuals on an active dietary intervention to an observational cohort introduces potential biases. The controlled intervention induces a specific metabolic response under tightly regulated conditions (e.g., consistent adherence, fasting state, and controlled macronutrient intake), whereas individuals in the observational cohort may exhibit similar metabolic profiles due to diverse, uncontrolled factors—such as underlying health conditions, medication use, or lifestyle behaviors—that are unrelated to intentional dietary changes. This disconnect may confound the association between diet-like metabolomic signatures and breast cancer risk

Within the controlled trials, key limitations include 1) the relatively small sample size, which limits statistical power and thus the ability to detect subtle diet-induced metabolic changes, 2) the short duration of these interventions (4-6 weeks), which may not fully capture long-term metabolic adaptations or their potential influence on breast cancer risk, and 3) lack of control of baseline metabolite and lipid levels of participants in the dietary intervention studies, which may have affected participants’ response to the interventions themselves^56^. Moreover, the metabolomic score used in this study is derived from a linear combination of metabolites, which may oversimplify complex metabolic pathways and potentially misrepresent underlying biological mechanisms.

In the NHSII cohort, a major limitation is the lack of applicability of these findings to postmenopausal populations. Because the NHSII cohort was predominantly premenopausal at the time of blood collection, the applicability of these findings to postmenopausal populations— who represent a substantial proportion of breast cancer cases—remains uncertain. Future studies should investigate whether similar metabolic mechanisms contribute to postmenopausal breast cancer risk. Further, fasting status was not uniform across participants, with higher fasting rates observed in the highest quartiles of both KD and LFD scores. However, the association between metabolomic scores and breast cancer risk remained significant when restricted to fasted individuals (**Supplementary Table 4**).

While our findings suggest plausible biological mechanisms, such as the role of CEs in steroidogenesis and the differential effects of TG species on cancer metabolism, the specific mechanism by which the metabolic changes induced by KD and LFD influence breast cancer remains unclear. Residual confounding is also a concern, as factors like genetic predisposition and unmeasured metabolites or hormones may contribute to the observed associations. Additionally, the nested case-control design allows for the identification of associations but does not establish causality between the metabolomic score and breast cancer risk. Future research incorporating larger, longer-term studies with randomized designs will be essential to validate these findings and further elucidate the causal mechanisms linking diet to breast cancer risk.

## 5 Conclusion

This study demonstrates that hypocaloric ketogenic and low-fat diets, despite differing macronutrient compositions, induce shared metabolic changes that are significantly positively associated with breast cancer risk in a premenopausal population. As hypocaloric diets reduce adiposity and, in the case of the KD, decrease circulating insulin levels, this positive association was unexpected.

The positive association between diet-specific metabolomic scores and breast cancer risk underscores the complexity of metabolic reprogramming in weight-loss interventions and the critical nature of dietary lipid intake during hypocaloric states. Our findings highlight the importance of understanding the metabolic consequences of dietary interventions, particularly for cancer prevention strategies. Future research should prioritize elucidating the mechanistic underpinnings of these associations as well as the specific lipid species involved in avoiding potentially detrimental effects of hypocaloric-like states. It will be critical for future studies to expand investigations to more diverse populations, including postmenopausal women, to refine dietary recommendations for cancer prevention.

## Data Availability

The data used in this study are available from the Nurses' Health Study but are not publicly available due to participant privacy and informed consent restrictions. Researchers may request access to the data by submitting a proposal to the Nurses' Health Study Steering Committee. Approval is subject to review and compliance with data use agreements and ethical guidelines.

https://github.com/krumsieklab/hypocaloric-diet-bc

## Acknowledgements

We would like to thank the participants and staff of the Nurses’ Health Study 2 for their valuable contributions. The authors would like to acknowledge the contribution to this study from central cancer registries supported through the Centers for Disease Control and Prevention’s National Program of Cancer Registries (NPCR) and/or the National Cancer Institute’s Surveillance, Epidemiology, and End Results (SEER) Program. Central registries may also be supported by state agencies, universities, and cancer centers. Participating central cancer registries include the following: Alabama, Alaska, Arizona, Arkansas, California, Colorado, Connecticut, Delaware, Florida, Georgia, Hawaii, Idaho, Indiana, Iowa, Kentucky, Louisiana, Massachusetts, Maine, Maryland, Michigan, Mississippi, Montana, Nebraska, Nevada, New Hampshire, New Jersey, New Mexico, New York, North Carolina, North Dakota, Ohio, Oklahoma, Oregon, Pennsylvania, Puerto Rico, Rhode Island, Seattle SEER Registry, South Carolina, Tennessee, Texas, Utah, Virginia, West Virginia, Wyoming.

## Funding

This study was funded by the National Institutes of Health (NIH)/National Cancer Institute (NCI) with the following grants: UM1 CA186107, P01 CA87969, R01 CA49449, R01 CA050385, U01 CA176726, U01 CA260352, R01 CA067262 and Susan G. Komen Foundation.

Funding for this study also came from 2020 AACR-The Mark Foundation for Cancer Research “Science of the Patient” (SOP) Grant (20-60-51-GONC), NIH R01CA279561, and Cycle for Survival.

## Disclosures

M.D.G. holds equity in Faeth Therapeutics and Skye Biosciences; reports consulting or advisory roles with Almac Discovery, Genentech Inc, Faeth Therapeutics, Scorpion Therapeutics, and Skye Biosciences; honoraria from Pfizer Inc.; patents, royalties, and other intellectual property with Weill Cornell Medicine and Faeth Therapeutics.

V.M. has institutional grants or contracts with AstraZeneca, Bristol Myers Squibb, Cullinan Oncology, DualityBio, Eisai, Faeth Therapeutics, Karyopharm Therapeutics, Merck, Takeda, and Zymeworks; receives personal meeting/travel support from AstraZeneca; and is an unpaid consultant for Clovis Oncology, Cullinan Oncology, DualityBio, Eisai, Faeth Therapeutics, Jazz Pharmaceuticals, GlaxoSmithKline, Immunocore, iTeos Therapeutics, Karyopharm Therapeutics, Lilly, Merck, Mereo BioPharma, MorphoSys, MSD, Novartis, Regeneron, Sutro Biopharma, and Zymeworks.

JK is co-founder and holds equity in iollo and ExactRx and is an advisor to Everfur (Stand Health Inc).

## Source Code

Source code for all analyses and figures of this study is available at https://github.com/krumsieklab/hypocaloric-diet-bc.

## Notes

### Author Declarations

IRB of Brigham and Women's Hospital gave ethical approval for this work IRB of Harvard T.H. Chan School of Public Health gave ethical approval for this work

