## Supplementary Tables and Figures for "Metabolomic signatures of hypocaloric dietary interventions associate with breast cancer risk in the Nurses’ Health Study II"

Supplementary Table 1: OSU Cohort Dietary Composition (taken from Buga et al, 2021).

|  | KD + Ketone Salts* | KD + Placebo* | LFD* |
| --- | --- | --- | --- |
| Energy (kcal/day) | 1845 ± 102 | 1752 ± 98 | 1900 ± 102 |
| Protein (g) | 99 ± 3 | 100 ± 3 | 100 ± 3 |
| Carbohydrate (g) | 40 ± 8 | 38 ± 7 | 259 ± 8 |
| Sugar (g) | 17 ± 3 | 17 ± 3 | 101 ± 3 |
| Fiber (g) | 10 ± 1 | 10 ± 1 | 34 ± 1 |
| Added sugars (g) | N/A | N/A | < 25 g/day |
| Fat (g) | 143 ± 9 | 131 ± 8 | 51 ± 9 |
| SFA (g) | 63 ± 4 | 63 ± 4 | 17 ± 4 |
| MUFA (g) | 38 ± 3 | 38 ± | 10 ± 3 |
| PUFA (g) | 8 ± 1 | 8 ± 1 | 7 ± 1 |
| Sodium (mg) | 6100 ± 32 | 2351 ± 30 | 1974 ± 31 |
| Potassium (mg) | 2211 ± 73 | 2243 ± 75 | 2758 ± 78 |
| Calcium (mg) | 2001 ± 36 | 880 ± 34 | 1008 ± 35 |

* All measurements given in mean ± SEM

Supplementary Table 2: MSK Cohort Dietary Composition (taken from Dantas et al, 2025).

| **Average Daily Nutrient Composition** | **AVG** | **SEM** |
| --- | --- | --- |
| Cals (kcal) | 2139 | 110 |
| Energy Density (kcal g-1) | 1.9 | 0.1 |
| Carbohydrate (%)^1^ | 6.2% | 0.0 |
| Total Fiber (g 1,000 kcal-1) | 6.9 | 0.2 |
| Sugar (g 1,000 kcal-1) | 4.5 | 0.4 |
| Protein (%)^1^ | 8.7% | 0.0 |
| Fat (%)^1^ | 87.2% | 0.0 |
| SFA (g 1,000 kcal-1) | 45.6 | 0.7 |
| MUFA (g 1,000 kcal-1) | 19.5 | 1.1 |
| PUFA (g 1,000 kcal-1) | 8.3 | 0.6 |
| Omega3 (g 1,000 kcal-1)^2^ | 0.8 | 0.1 |
| Omega6 (g 1,000 kcal-1)^2^ | 5.3 | 0.5 |
| Sodium (mg 1,000 kcal-1) | 788.7 | 19.6 |

^1^ Energy attributable to each macronutrient was calculated using Atwater factors (protein and carbohydrate each 4 kcal/g, fat 9 kcal/g)

^2^ Tabulated values are an underestimate of true intake as nutrient composition data for n-3 and n-6 fatty acids were missing for 20% of ingredients. The proportion of ingredients per menu missing n-3 and n-6 composition values were 20.3% (SD 1.49%).

Supplementary Figure 1: Diet scores on controlled trial participants over weeks of intervention

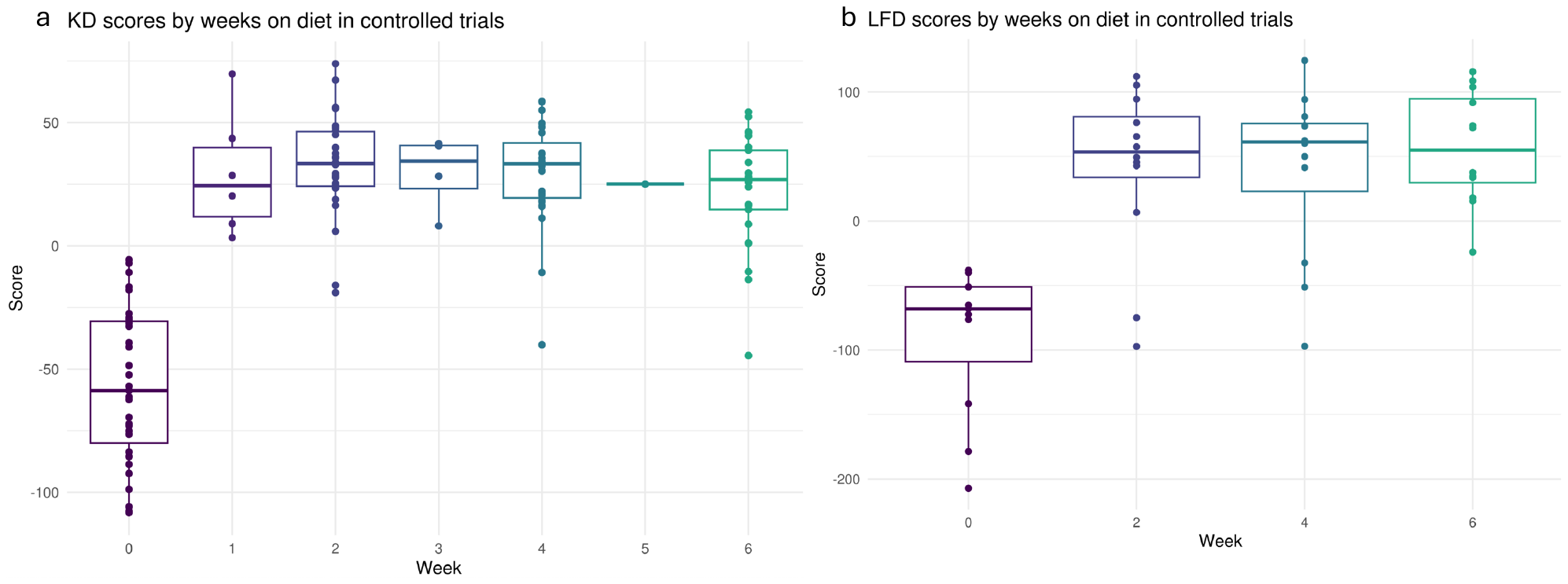

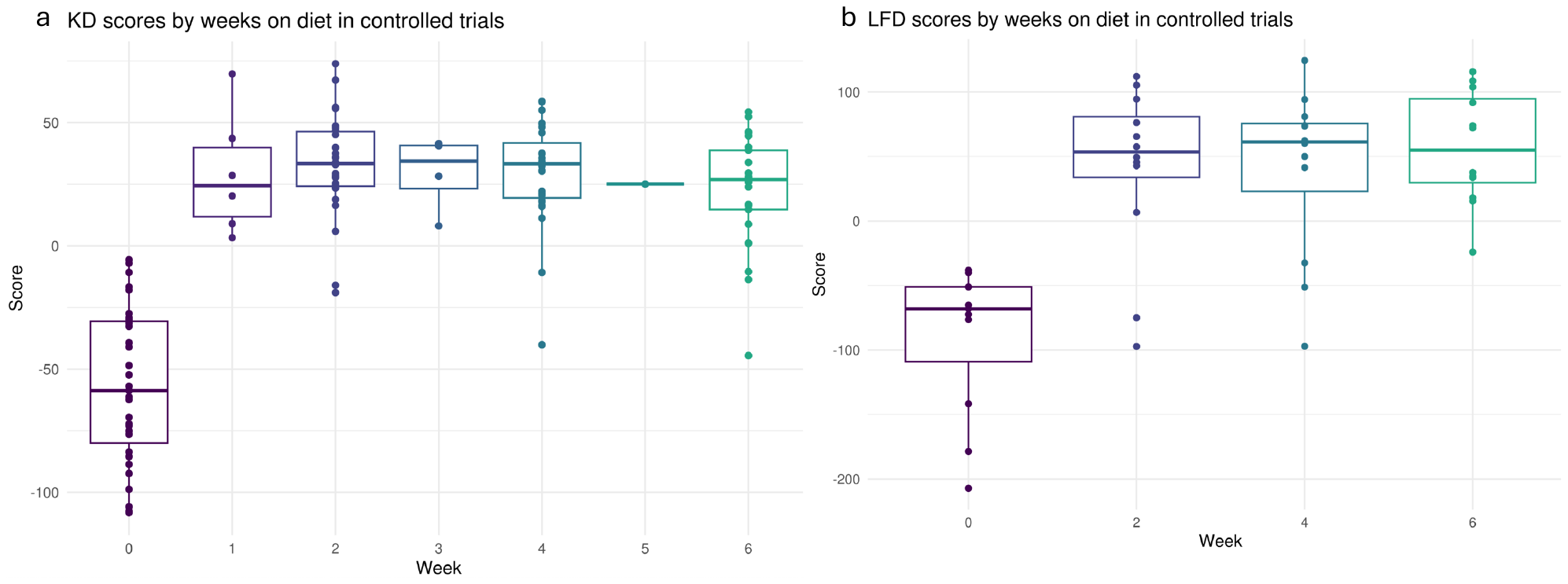

Supplementary Table 3: Descriptive characteristics of Nurses’ Health Study II participants tabulated by Low-fat Diet metabolomics score quartiles among controls (N = 1054)

| Characteristic | Quartile1  N = 264  ( < -41.53) | Quartile 2  N = 263  ( $\boldsymbol{\geq}$ -41.53to < -0.67) | Quartile 3  N = 263  ($\boldsymbol{\geq}$-0.67 to < 37.16) | Quartile 4  N = 264  ( $\boldsymbol{\geq}$ 37.16) |
| --- | --- | --- | --- | --- |
| Age at blood draw (Years), mean (SD) | 46.4 (4.1) | 45.3 (4.2) | 44.3 (4.2) | 43.3 (4.6) |
| Fasting at blood draw (>8 hours), N (%) | 187 (71%) | 202 (77%) | 195 (74%) | 206 (78%) |
| Menopausal status and PMH use at blood draw, N (%) |  |  |  |  |
| Postmenopausal – no hormone use | 10 (3.8%) | 3 (1.1%) | 6 (2.3%) | 4 (1.5%) |
| Postmenopausal – yes hormone use | 43 (16%) | 28 (11%) | 19 (7.2%) | 25 (9.5%) |
| Premenopausal | 172 (65%) | 200 (76%) | 214 (81%) | 223 (84%) |
| Unknown | 39 (15%) | 32 (12%) | 24 (9.1%) | 12 (4.5%) |
| Age at Menarche (Years), mean (SD) | 12.3 (1.5) | 12.4 (1.4) | 12.7 (1.4) | 12.5 (1.4) |
| Nulliparous, N (%) | 43 (16%) | 48 (18%) | 48 (18%) | 49 (19%) |
| Parity, mean (SD) | 2.4 (1) | 2.4 (1) | 2.3 (1) | 2.3 (0.9) |
| History of benign breast disease, N (%) | 47 (18%) | 41 (16%) | 38 (14%) | 38 (14%) |
| Family history of breast cancer, N (%) | 38 (14%) | 36 (14%) | 16 (6.1%) | 24 (9.1%) |
| BMI at age 18 (kg/m^2^), mean (SD) | 21.7 (3.3) | 20.9 (2.9) | 20.9 (3.1) | 21.1 (2.9) |
| Weight change since age 18 (kg), mean (SD) | 20 (15) | 13 (11) | 10 (10) | 9 (11) |
| Activity levels at blood draw (MET-hours/week), mean (SD) | 18 (29) | 17 (18) | 20 (22) | 18 (19) |
| Current Smoker, N (%) | 23 (8.7%) | 20 (7.6%) | 13 (4.9%) | 18 (6.8%) |
| Alcohol Consumption at blood draw (g/day), mean (SD) | 2.7 (5) | 3.7 (7) | 3.9 (6.8) | 2.9 (5.2) |

Supplementary Table 4: Odds ratios and 95% Confidence Intervals for quartiles of ketogenic diet metabolomics scores and breast cancer risk in Nurses’ Health Study II fasted samples only (725 cases/790 controls)

| **Ketogenic Diet Metabolomics Score** | **Categorical OR (95% CI)** | | | | | **Continuous OR (95% CI)** | |
| --- | --- | --- | --- | --- | --- | --- | --- |
| **Overall breast cancer (725 cases)** | Q1 | Q2 | Q3 | Q4 | P-trend^4^ | Per SD increase | P-value |
| Multivariable adjusted^1^ | 1.0 (ref) | 1.21 (0.87, 1.67) | 1.22 (0.89, 1.66) | 1.39 (1.01, 1.91) | 0.06 | 1.09 (0.98, 1.22) | 0.118 |
| Multivariable adjusted + BMI^2^ | 1.0 (ref) | 1.19 (0.86, 1.65) | 1.2 (0.87, 1.63) | 1.37 (0.98, 1.9) | 0.07 | 1.09 (0.97, 1.22) | 0.146 |

**1** conditional logistic regression adjusted for smoking, alcohol intake, physical activity, age at menarche, parity and age of first birth, family history of breast cancer, and personal history of benign breast disease

**2** additionally adjusted for BMI at age 18 and weight change since age 18 to blood draw.

Supplementary Table 5: Odds ratios and 95% Confidence Intervals for quartiles of low-fat diet metabolomics scores and breast cancer risk in Nurses’ Health Study II fasted samples only (725 cases/790 controls)

| **Low-fat Diet Metabolomics Score** | **Categorical OR (95% CI)** | | | | | **Continuous OR (95% CI)** | |
| --- | --- | --- | --- | --- | --- | --- | --- |
| **Overall breast cancer (725 cases)** | Q1 | Q2 | Q3 | Q4 | P-trend^4^ | Per SD increase | P-value |
| Multivariable adjusted^1^ | 1.0 (ref) | 0.99 (0.71, 1.38) | 1.44 (1.05, 1.99) | 1.37 (1, 1.89) | 0.01 | 1.11 (1, 1.25) | 0.059 |
| Multivariable adjusted + BMI^2^ | 1.0 (ref) | 0.98 (0.69, 1.37) | 1.41 (1.01, 1.96) | 1.34 (0.96, 1.86) | 0.02 | 1.1 (0.98, 1.24) | 0.099 |

**1** conditional logistic regression adjusted for smoking, alcohol intake, physical activity, age at menarche, parity and age of first birth, family history of breast cancer, and personal history of benign breast disease

**2** additionally adjusted for BMI at age 18 and weight change since age 18 to blood draw.
